# Factors influencing the adoption of health education for Schistosomiasis control in Luangwa, Zambia

**DOI:** 10.1101/2024.06.20.24305247

**Authors:** Rabecca Chinoya, Professor Joseph Zulu, Dr Hikabansa Halwiindi, Mr Adam Silumbwe, Ms Patricia Maritim

## Abstract

Implementation of health education with mass drug administration is advised so as to induce better decision among various populations through uptake of the health messages. However, this disease remains a problem in Luangwa despite implementation of health education. The study was thus conducted to assess the factors influencing the adoption of health education for schistosomiasis control in Luangwa district of Zambia. It was specifically aimed at assessing the scope of health education and strategies being employed in delivery of health education for schistosomiasis in Luangwa. Additionally, it explored the barriers and facilitators in delivering health education for schistosomiasis in Luangwa and finally revealed perceptions of key stakeholders towards health education messages for schistosomiasis in Luangwa.

This case study qualitatively explored various factors influencing adoption of health education in Luangwa district. Three Key Informant Interviews, six In-depth Interviews and eight Focus Group Discussions were performed with implementers, health educators and recipients of the health education respectively. The recipients of the health education included school going children, members of the community and mothers who attended under-five clinic sessions.

The Factors that were seen to facilitate acceptability of health education include adaptability of health education, community engagement, use of locally acceptable venues to deliver health education, use of teaching aids and learning from others. On the other hand, factors that inhibited acceptability of health education were poor road network, unreached areas, beliefs and myths surrounding bilharzia, the fishing cultural orientation, inadequate income and human resources, busy work schedules and poor access to clean water.

The framework used in the study was found to reveal critical aspects of Health education adoption in Luangwa district. However, there still exists a need to conduct further research as well as focus on improving the structural characteristics as are illustrated in this study.

## Background

Schistosomiasis (bilharzia) is a neglected tropical disease (NTD) caused by blood flukes belonging to the phylum Platyhelminthes and genus *Schistosoma.* It has been prioritized by World Health Organization (WHO) for control and possible elimination by 2020 (Lothe et al., 2018). Morbidity of schistosomiasis is attributed to 3 species namely: *Schistosoma mansoni, Schistosoma haematobium* and *Schistosoma japonicum* (Tuhebwe et al., 2015). A global estimate of 240 million people are infected, while over 700 million are estimated as being at risk of in 78 developing countries of the world. (Tuhebwe et al., 2015) with a global estimate of 70 million disability adjusted life years lost annually (Odongkara et al., 2006). In Sub-Saharan Africa, approximately 25% of the total population is infected with schistosomiasis (Lothe et al., 2018) and Zambia’s prevalence of schistosomiasis is highest in poor communities thus showing a need for health education as a preventive measure against schistosomiasis (Kalinda et al., 2018).

Mass drug administration (MDA), which is Preventive chemotherapy (PC) remains the number one strategy endorsed by WHO (World Health Organisation) for the prevention and control of schistosomiasis. Schools regularly treat school going children for schistosomiasis, through coordinated use of reasonably priced drugs (Deng et al., 2018). WHO encouraged the need to adopt programs that focus on elimination rather than just control of disease burden (Organization, 2010). In addition to PC, hygienic and sanitation interventions, which includes health education are required if the transmission of schistosomiasis is to be interrupted (Sokolow et al., 2018).

Whilst there is recorded progress of the implementation of the MDA the challenges in other settings, mostly Low- and Middle-Income Countries like Zambia still exist, as schistosomiasis burden remains a challenge (Kalinda et al., 2018). This then can follow with the views of Tchuem Tchuente et al. (2017) who emphasized the need for modification, adaptation and change of strategies towards elimination of schistosomiasis by explaining that, the disease had concomitantly raised new challenges, which required consideration. Furthermore, it was explained that most strategies employed were strictly for the control of morbidity and were employed at a time when praziquantel, a drug used in preventive chemotherapy, was scarce (Tchuem Tchuente et al., 2017).

Mass drug administration alone has been seen to not be effective without modification of personal hygiene which is achieved through integration of health education (Qian et al., 2019) which has been neglected in the time of prioritising MDA alone (Ross et al., 2017). The integration of health education is expected to promote behaviour change and in turn ensure sustainability of MDA and compliance to the preventive drug (Qian et al., 2019).

As stated by (WHO, 1990), the consolidation of behaviour change only becomes visible through reduction of disease burden. However, this has not been the case with Luangwa district even after implementation of health education in various settings such as schools, clinics and communities. This led to the question of whether the health education messages are being taken up by the targeted population as it has proved to be effective for the control of schistosomiasis in other settings such as China (Tchuem Tchuente et al., 2017).

In his study on the assessment of health education, Lansdown et al. (2002) showed that there was a need to incorporate various activities in delivering HE and there was a need for the Health educators to teach what they considered more relevant to the beneficiaries. The little evidence that exists shows little integration of HE education into routine practice and therefore evidence of intervention benefits is lacking (Fleming et al., 2009). It was further mentioned by the beneficiaries that HE messages were only delivered on days of MDA delivery and this was not very effective (Fleming et al., 2009).

A recommendation was established by Rassi et al. (2016) that focus should be on teaching about the causes of the disease, ways of transmission, prevention and treatment rather than awareness of the disease. Some of the forms of delivery of Health education that proved successful were Graphical design products, Daily-use products, audio-visual products and Comprehensive (combination of two or all three of the products) products (Qian et al., 2019). Engels et al. (2002) encouraged the need to involve other strategies such as provision of safe water and adequate sanitation, which aid in taking up health education messages. In a study conducted in Kenya by Mwai et al. (2016) , one of the participants mentioned that door to door visits were ideal because people already knew each other and they could easily do it on their own.

In a study in Tanzania, the target was the primary school children in order to improve their personal hygiene with respect to hand washing after using the latrine, keeping school environments and the latrines clean, having footwear, using latrines and being aware of the risks of contact with contaminated water. Some of the observations made at the schools were the classroom teachings, which was generally on personal hygiene but included light components on the need for clean and safe water as well as plays and sketches and incorporation into subjects such as geography, KiSwahili and science (Lansdown et al., 2002). In a study to assess knowledge and perceptions of people of Nampula in Mozambique, concluded that the lack of adequate knowledge on transmission and preventive measures and existing misconceptions about schistosomiasis created a barrier towards successful disease prevention and control (Rassi et al., 2016). Individual characteristics are therefore necessary in understanding the adoption of these health education messages (Damschroder et al., 2009).

It is clear that comprehensive reports/ writings have been documented to show barriers to adopting Health education on various constructs with majority on individual characteristics but less compilations have been made with the domains of the consolidated framework and in particular the constructs to be documented in this study.

## Methods

The study was conducted in Luangwa, a district in Lusaka province of Zambia where MDA and health education have been implemented with incidence rate recently being at 20%. Luangwa is one of the districts in Zambia having high incidence rates of bilharzia. The district is approximately 3,872 km² with Density 7.620/km² and had a population of 24,304 at last census in 2010 (Central Statistical Office Zambia (web), 2018). This district is considered rural and most of its economic activity being around fishing which is a key factor in schistosomiasis transmission. The study site was identified after a report of 2018 which showed an increasing incidence of schistosomiasis and the recruitment period for the study was from 3^rd^ March, 2020 to 12^th^ March, 2020. Purposive sampling was used and interviews were carried out with district health office (n=3), facility health workers & Community health workers (CHWs) (n=6), while pupils and community members participated in the FGDs (m=8). The key informant interviews (KIIs) and In-depth interviews (IDIs) were conducted with the aid of two different interview guides and FGDs were conducted with the aid of focus group discussion guides. The key informant interviews were intended to seek information on the implementation of the health education, which included, the topics covered, the strategies used as well as the challenges faced during implementation of health education. The in-depth interviews sought to explore the various issues surrounding delivery of HE and the FGDs were used to acquire information on what drove decision making for people in Luangwa to either adopt or not take up the HE.

A thematic analysis approach was used to identify and analyse patterns within the data that had been collected. With reference to the key questions that were to be discussed and the already available information on adoption of health education in different settings as well as information of evaluation of interventions according to the consolidated framework. A coding list was generated and theme development was according to responses provided by the participants and reduction of data was by noticing relevant phenomena, collecting examples of the phenomena, and analysing them to find similarities, differences, patterns and overlying structures.

## Ethics approval and consent to participate

The proposal of this study was submitted to the University of Zambia Biomedical Research Ethics Committee and Nation Health Research Authority. Both institutions are situated in Lusaka, Zambia and both approved conduct of this research. Due to the study being health related and involving pupils in various schools, approval was also sought and obtained from Provincial Health Office in Lusaka, Provincial Education Office in Lusaka, District Health Office in Luangwa and District Education Board in Luangwa. The study gave a choice to those who were willing to assist in revealing information regarding the factors of adoption an opportunity whether they wanted to participate in the study or not. The study was intended to provide maximum benefits for the population of Luangwa as an assessment of the adoption process would enhance its effective implementation and hence possibly throw a positive arrow towards the incidence reduction. Generally, Participation in the survey was voluntary, confidentiality and anonymity were ensured, and written informed consent was sought from all interviewees.

## Results

The table below (*Table 2*) shows the various themes arising from the data collected as well as the various codes under specific themes. It also shows the relation of the data collected to the various domains of the Consolidated Framework for Implementation Research. The IDs were maintained in the manuscript as they were and are not known to anyone outside the research group.

**Table 1:** incidence rates in Luangwa from 2015 to 2018

| Year | Population (incidence) |  |  |
| --- | --- | --- | --- |
|  | Under 5 years | Above 5 years | ALL |
| **2015** | 5591<br>(6.8%) | 22363<br>(7.11%) | 27954<br>(7.05%) |
| **2016** | 5685<br>(10.71%) | 22782<br>(13.83%) | 28477<br>(13.20%) |
| **2017** | 4820<br>(15.35%) | 24146<br>(15.24%) | 28966<br>(15.26%) |
| **2018** | 4910<br>(22.81%) | 24596<br>(19.52%) | 29506<br>(20.06%) |

Information adopted from a report by Luangwa District Health Office

**Table 2:**
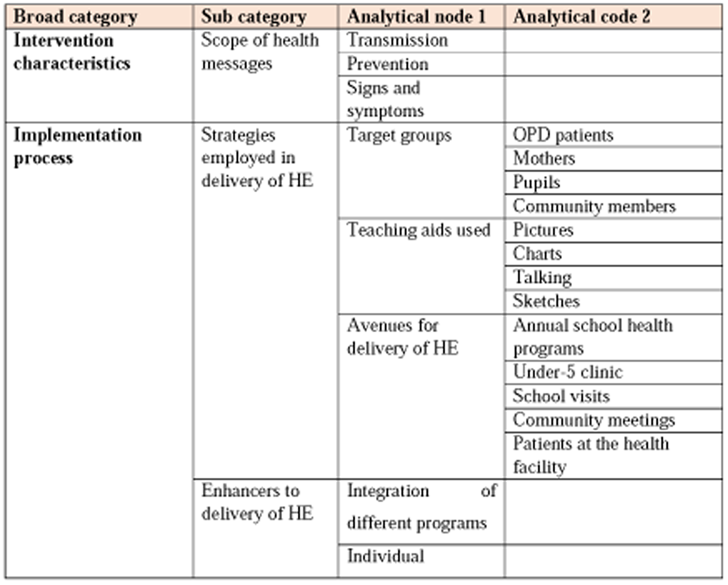
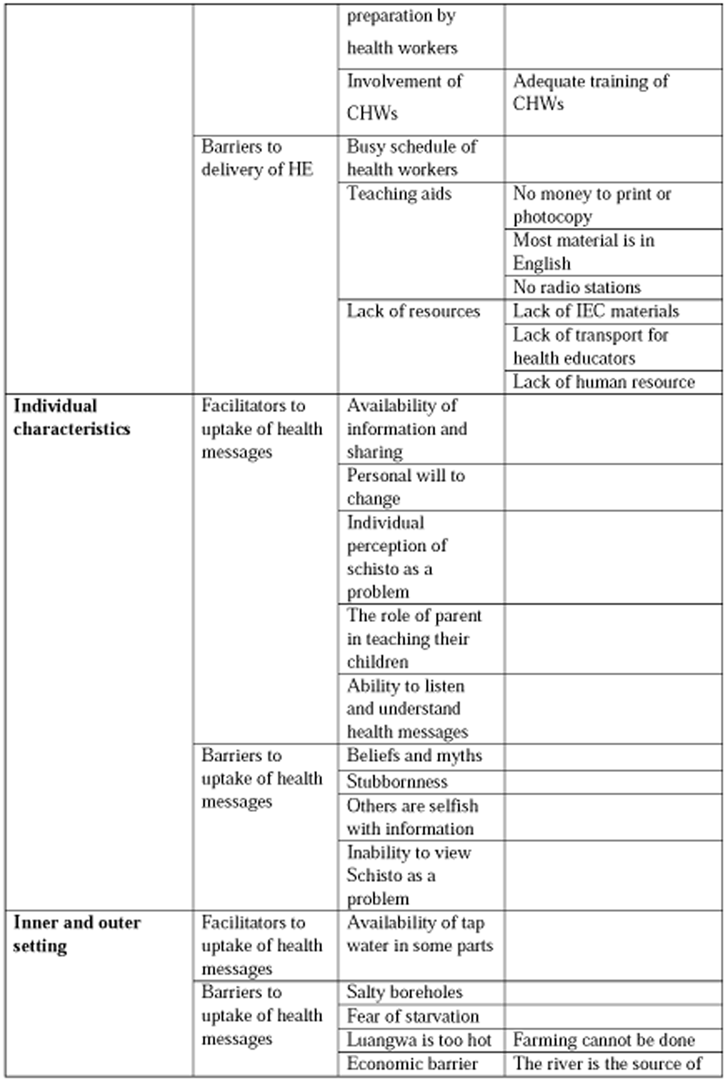
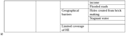
Qualitative data analysis code-list

### Adaptability of health education

Health education for schistosomiasis in Luangwa has various topics which include definition of schistosomiasis, risks & transmission, signs and symptoms and prevention & control with treatment not so often taught as they aim only to impart knowledge regarding the prevention. In Luangwa district, the health educators made it easy for the recipients to understand the topics by leaving out the life cycle of schistosomiasis. This was because they felt it would be easier for the illiterate target populations to understand.

*“…mainly we give uhm, we start with the risks that are associated or how one can get bilharzia and the factors that may lead to bilharzia…we will tell them how bilharzia comes about; you can’t really go in details of the life cycle but you will tell them that it’s attached to the water and the likes. Yeah, so you tell them how it comes about prevention and subsequently treatment though treatment is silent because our aim is to impart knowledge on the preventive part…” [KII1_Health worker]*

One of the health educators, a midwife, mentioned that she first defined what bilharzia was and would in most cases realize that they still remembered as they would respond <u>‘kutunda</u> <u>blood’</u> (releasing blood when urinating). She would then go further into educating them on the life cycle of bilharzia and would encourage them to wear boots as they went to the river to do their washing and also told them to avoid swimming in the rivers and finally educate them on the signs and symptoms.

*“…I was just defining what bilharzia is and you would find even they know it; you would ask them what is bilharzia? They will answer you ‘kutunda blood’.” [IDI2_Midwife]*

In reviewing success of health education in Luangwa, it was evident because majority of the participants were able to explain what schistosomiasis was and how it was caused. However, a minor population of people were ignorant of the cause of schistosomiasis.

*“…stagnant water has germs which can even get into nails when one steps in the water and then gain entry into the body.” [FGD2-8_ community member]*

*“…bilharzia can come from contaminated food such as left overs which contain germs” [FGD2-3_Community member]*.

Many people in Luangwa depend on market gardening and this is done in areas that are water logged and this is evidence of danger to them as they have constant interaction with the water infested with parasites. The gardeners are told that when they notice blood in their urine, they ought to go to the health facility as quickly as possible so that case management can be done. Most children go to the river to do their washing and bathing and are taught to avoid urinating in the water as this would lead to the spread of bilharzia in Luangwa.

*“…they teach us about prevention as well as signs to signify one with bilharzia. Sometime back…” [FGD4_pupils]*

### Community engagement

The use of community volunteers aids the delivery of the health messages for schistosomiasis because the language barrier is partly broken and they are closer to the target population thus enhancing success of health education. With the use of community health workers, there is no need to worry about transportation as they are within a walkable distance from their homes or health facility.

*“…uhm, because looking at our situation, here, our health workers cannot manage to educate the community, like going out there. So, what is done is community health workers are selected, there are groups in different villages, so, those are educated and sent deeper into the villages and they educate…” [KII2_Health worker]*.

The CHWs are also adequately trained to ensure they offer relevant information to the various populations. The health workers ensure that the messages given to the CHWs and CBVs are designed from the central level as well as from the district level so that the messages given to the community are uniform and straight to the point. Another activity they have is the TS-technical support as well as mentorship where they are given the messages required to be delivered in the community.

The role of parents in teaching and warning their children in homes was also one of the enhancers to uptake of HE as children saw their parents as leading examples. The ability to listen and understand the messages by various individuals also led to uptake of the HE, most of whom were taught during their days in primary school.

*“… I used to play in the drainages with my friends on our way to the market after it rained. One day, someone told me not to be walking or playing in the drainages because there were some diseases caused by rain water, I ignored and continued, then on another day my mum told me that there was a disease caused by stagnant water and advised me to stop playing in such water and I stopped.…” [FGD5-6_Pupil]*.

### Use of locally acceptable venues to deliver health education

There are various groups targeted for health education such as OPD patients, Mothers who go for under 5 clinics, Pupils and the community members and various avenues are used and sometimes taken advantage of for the delivery of HE. One of the avenues being the Annual School health program which involves few school visits to teach pupils. The mothers are taught during Under-five clinic sessions which is a way of integration of various health programs while community meetings are used to reach the members of several communities/ villages.

*“…before patients tell us what’s wrong, like 30 minutes before we start that, we educate them on different things…” [IDI1_Clinical officer]*

*“…Yes, for the mothers, I would educate them also but I would concentrate more on the pupils…” [IDI2_Midwife)*

### Use of teaching aids

The teaching aids used for bilharzia have information of a life cycle, symptoms & prevention. Among the teaching aids used were pictures, handouts, charts & drama in various avenues which were intended to impart deep understanding on bilharzia in view of enhancing social behavioural change. Explanations or talking stood as the most used form of teaching as there was lack of resources to aid the production i.e., photocopying or printing of IEC materials. Other times when pictures and brochures were available, in English and Nyanja, the members of various communities mentioned that they had been given the brochures to carry home as well as pictures showing worm infestations to go and stick in their homes.

*“…We have teaching aids that we use on bilharzia, which has that information of a life cycle, symptoms and prevention part, so we take advantage of going through schools where there are people and we also when we get chance teach at the clinics where people come. Then the main avenue we use are schools because a lot of them are clustered in that places.” [KII1-Health worker]*

### Learning from others

Most of these were people who had suffered from bilharzia and others being those who saw others, be it their relatives or neighbours suffer from the disease. Some of the people did not have to see or experience it first-hand but were able to understand bilharzia as a problem through the various teachings given to them. All this is encompassed in a Personal-will to change as none of the individuals mentioned being forced to change their behaviour.

*“… yes, I suffered from bilharzia when I was between 12 and 14 years old, but I quickly went to the health facility and was given very big tablets and I drank them all. I fear it coming back but I know it may not come back because I now protect myself. When I go in search of fish, I sit in my boat, I don’t step in the water, I only throw my rod and when it comes to bathing, I use water from the borehole” [FGD6-1_Pupil]*

## Factors inhibiting acceptability of health education

### Poor road network

One other factor that inhibited uptake of HE was the flooding of the main road during the rainy season. This is the road certain villages such as Kamoba can use to get to the market because the short cut always has stagnant water and pose a major risk towards schistosomiasis. However, some people mentioned that they could not use the main road as it took them a much longer time to get to the market and sometimes had elephants present. Other than the adults, school children were left without a choice as they would come back tired from school, hence opting to use the shortcut with stagnant water.

*“…the shortcut to the BOMA market is through the same contaminated water because the whole road gets flooded when it rains.” [FGD2-8_Community member]*

### Some areas not reached

The lack of information was observed to be one of the barriers to uptake of HE as it was mentioned that the people of Chidada and Indeco (communities) were lacking information on bilharzia prevention as the two communities are populated by elderly people who are only given food and provided shelter. In Katondwe area, the members of the community expressed concern on how they had not been receiving bilharzia health messages for many years and called for the commencement of sensitization in the area.

### Beliefs and myths surrounding bilharzia

Normalisation of unhealthy behaviour among the young people interviewed through this study was seen to be a hinderance to adoption of health education. It was observed that majority of them complained about it being too hot and this led them to go to the river for the purpose of cooling themselves.

*“…I can’t stop going to the river because we live at the corner and it is very hot, especially in October, so, there’s nothing we can do because even in class, we can’t wait to knock off so that we go to the river…” [FGD6-4_Pupil]*

Some of the beliefs and myths surrounding bilharzia and its strategies of prevention have led to the lack of uptake of HE among certain individuals. A middle aged woman of Katondwe revealed that some members of the community chose to not pay attention to the bilharzia messages as they believed bilharzia was caused by neighbours who had previously made threats on others. Some of the individuals believed that bilharzia was a curse and not a disease that could come from their interaction with water.

“…*some people have been taught about bilharzia and its causes, even so, they continue to go and swim and when they release blood during urination, they say, ‘I have been cursed by my great great great grandma.’ That is what they say…” [FGD5-6_Pupil]*

#### The finishing cultural orientation

The introduction of clean water had prevented some from going to fetch water at the river but are forced to go there for the purpose of looking for relish. Luangwa district is known for its fish, a number of people brought up the point of having to step in stagnant water when one goes to fish and this puts them at risk of contracting schistosomiasis. This causes some people to not change their behaviour as they will mention that fishing is their only way of sustaining themselves and telling them to not go to the river shows that the educator is against their only source of food.

“*You can tell them or try to advise them on their interaction with water and they will say ‘you don’t provide for me’ and as a child, I’m hurt by such words and its scary to continue teaching them” [FGD6-4_Pupil]*.

Majority of the population brought out the fact that Farming cannot be done due to the geography of the district and this causes them to solely depend on fishing.

*“…I still go to the river because we need relish and this place has no farmers to provide a variety of relish, so, we choose to go and fish…” [FGD6-1_Pupil]*

Unwillingness to learn was another hinderance to uptake of HE as most people did not want to learn from the young ones, and even when they did learn, they did not take the messages with the seriousness they deserved as they preferred to continue in their ways. This was attributed by many as being due to failure to view bilharzia as a major problem.

*“…others change while others don’t. The reasons why people tend not to change is due to their attitude, they just don’t care to listen and ignore the teachings given to them. Mostly these are women and children, we have children whom we can tell about preventing bilharzia, but they will still go back to the source of the disease…” [FGD3-3_Mother]*

### Inadequate income

The lack of finances made it very difficult to effectively deliver health education messages in Luangwa district due to Lack of MDA sustainability which branches from lack of support for bilharzia specifically from the ministry and lack of partners such as NGOs to come on board and offer assistance. The lack of support from the ministry of health has been attributed to the discontinuation of the MDA as at a time of MDA, there was such support from the ministry and this aided delivery of health education to the people of Luangwa. This is due to bilharzia not being recognized as the major problem in Luangwa district as there are other diseases such as malaria and respiratory infections currently at the top of the list. The lack of radio station in the area was also called out as one of the barriers to delivery of health messages and having one would be making it easier to communicate and reach a wide range of the population.

*“…we don’t really have things like radio stations and all those things so, it is a bit difficult to use the other forms of media…” [KII2_Health worker]*

Lack of funds trickled to not being able to assist health educators with transport so that they could visit faraway places as well as lack of allowance for their extra hours of work. Some of the health educators mentioned that they were willing to go and teach but transportation was a limiting factor.

“*They are not enough, there’s you see, like I can only talk to certain households because time and maybe resources, I can’t reach certain communities because of transportation, and not supported at that particular time. It will take a long time for me to apply to the district office and ask for transport just for me to go and give health education on bilharzia……” [IDI6_EHT]*.

### Inadequate human resources

Human resource is one of the most important components in every organization and as such is an important component in the health sector for the delivery of health education messages. One of the EHTs made mention of how difficult it was for him as he was the only one available to teach at his place of work.

“*Here, we are quite limited when it comes to sensitizing because, mostly, I’m the only one who goes around, you have seen? quite alright I normally talk to our nurse there and we normally educate them when we have epidemic meetings, so that in a case that I’m on leave,*

*they should take part as well and sensitize. Mostly, if I’m not around, meaning health education pauses” [IDI5_EHT]*.

### Busy work schedules

With regards implementation, it was not easy for the health workers to incorporate health education into their routine work due to their busy schedules. Health education is a notable component of public health and as such EHTs together with other health workers such Clinical Officers and Nurses did participate in delivery of information to patients and the population at large. However, the health workers had other activities other than sensitisation and this causes them to only rush through the topics and it is difficult for them to teach on various topics due to lack of time.

*“…Time is a factor, so we just rush through… And there are other problems apart from bilharzia, so it can’t be bilharzia everyday…” [IDI1_Clinical Officer]*

### Limited sharing of information

However, the unwillingness by some individuals led to lack of sharing of information which was a hinderance to uptake of HE as those who learnt chose to not pass on the information because the people they would want to teach would say “*why are you telling me, you think I don’t know*?” Some pupils did not share information about Schistosomiasis because their parents had not been to school and this instilled fear in them as their parents would feel inferior and thus only preferred to share information when they were asked about what they learnt at school. Other pupils chose not to share information because of being scolded or told to not mention their school lessons by their friends who did not attend school.

*“… if we go in the community and chat with our friends, the moment you bring up a health education topic, those who do not go to school will threaten you and say’ stop talking about school things because this is not school’, so, in that way, you can’t be comfortable enough to teach them because they make it seem as though it is nonsense…” [FGD6-4_Pupil]*

### Poor access to clean water

Another challenge was the few taps in some parts of Soweto, a compound, which caused most people to prefer going to the river which was less crowded. One woman of Soweto complained about the payment required for one to drink water at the tap provided at the harbour, this does not sit well with them as they do not have money to pay for their drinking water when they go fishing and only have the option of drinking water directly from the river.

The Lusaka water and sewerage company has taken up the responsibility of making sure the people of Luangwa have clean water from their taps. Although not all villages have taps, the few villages that have been provided with this water have shown positive change as most of them now avoid going to the river for the purpose of fetching water to drink or for domestic use.

*“…with us who have taps in our homes, we no longer get water from the river…” [FGD1-11_Member of the community]*

The provision of clean water for the people of Luangwa has been implemented through provision of community and house hold taps as well as boreholes so as to limit people from visiting the river for their water. However, this has been received with mixed feelings as most of the people complained that some of the boreholes sunk in the communities are said to produce water containing salt and bicarbonate of soda.

*“…we have 4 boreholes and 2 of those boreholes produce salty water which also has soda which if we had to cook okra, it could cook very well and that is why we prefer water from the river. The other 2 are very far from our home and the river is much near. …” [FGD6-3_Pupil]*

## Discussion

With the aim of this study being to assess the factors influencing the adoption of health education for schistosomiasis control in Luangwa district its findings were found to be in line with studies conducted by Engels et al. (2002) and Hu et al. (2005). These studies suggested that in order to enable the effectiveness of HE, there was need for an integrated approach towards adoption of Health education in Luangwa district.

The messages given to the residents in Luangwa were simplified as is suggested by Rassi et al. (2016) who emphasized that only the necessary parts are taught as this makes it easier for the residents to understand. Majority of the study participants were able to explain the various aspects of schistosomiasis, however, some of them were still confusing it with diarrheal diseases. So, in as much as much of the population know about bilharzia, some still do not know despite being taught perhaps owing to the fact that they learn about various diseases at a time or they are not sure of what they learn about or are taught in tribes or languages they do not fully understand.

The various strategies employed towards the implementation of HE is necessary but it is evident that more ought to be done in order to facilitate the uptake of the health messages. It was necessary for the implementers to understand the target population and this was evident in Luangwa as the messages were kept simple so as to carter for the various population groups. Aside the reviewed studies, the implementers in Luangwa embraced various avenues and target groups which show how much the population of Luangwa was meant to be covered but some members of the district had still been left out and some of these are schools such as; Luangwa secondary school & Kapoche secondary school, communities such as Katondwe, Chidada & Soweto and mothers at under-5 sessions such as those at BOMA clinic.

Most of the Luangwa population that had been left out of the health education messages expressed their concern on how much they would love to have these health education sessions especially those from Kapoche secondary school and katondwe area. Some of the communities such as those in Soweto and Katondwe expressed concern over how they had not had HE sessions specifically for schistosomiasis for so many years while pleading that the teachings continue as were being done for malaria and HIV/AIDS. Some of the residents of Katondwe area mentioned the need to effectively implement door to door sensitization and this is necessary because in a study conducted by Mwai et al. (2016) who emphasized that people were familiar with each other and it could be done by volunteers in the various communities and the same thoughts were shared with the residents of Katondwe.

The use of various IEC materials is also a necessary step towards successful adoption of HE as various individuals have different levels of understanding. Drama was recommended by study participants as one of the best ways of delivering health messages as most people are easily attracted to the sounds of the drums and the sketches keep people entertained. As had been reported by Lansdown et al. (2002), visual aids such as pictures, billboards, posters and brochures were proposed as being very effective towards the delivery of health messages as they provided a primary knowledge on the transmission, treatment and control of schistosomiasis and fits well for all members of the community (Qian et al., 2019). Some pupils went on to mention that the use of pictures enabled those who were unable to read to adequately understand the messages through pictorial illustration regarding transmission as well as prevention.

The use of CHWs in the delivery of HE is a step in the positive direction as it is intended to break the language barrier as was mentioned by a key informant, but this has not been the case as the CHWs still use the regionalized language (Nyanja) to teach and this, to a certain degree defeats the purpose of bringing them in. The integration of various health programs has been helpful towards facilitating uptake as the recipients are able to have the HE amidst receiving other health services but the busy schedule of the health workers has proved as a disturbance towards this attainment. Besides the busy schedule of the health workers, the lack of resources has also been a barrier towards the uptake of the health messages as there is lack of IEC materials and health educators are not available because of lack of human resource and also lack of transport allowance to aid their movements to various destinations. All this has also led to reduced coverage of the HE as some schools and parts of some communities are not reached.

The availability of information and the possibility of information sharing also facilitated the uptake of HE as Rassi et al. (2016) suggested that the lack of information caused a barrier towards successful disease prevention. However, the inflexibility in those who received the information and those who were to be continuously shared with led to lack of uptake as some of them did not receive the information and others received the information but chose to ignore it. The unwillingness in some of the residents led to disruption of the will to listen and understand the messages and was hence a barrier towards uptake of the health education.

The individual perception of schistosomiasis as a problem by many was seen to facilitate uptake of health education as most of those who had an experience of it did not want to experience the painful schistosomiasis a second time. Others who did not even experience it themselves and saw it from their friends and family members shared the same feeling of not ever wanting to experience the disease. However, this personal will to change was distorted in some as they saw schistosomiasis as being a curse or resulting from witchcraft and this caused many of them to not follow the preventive measures laid down the health education delivery.

As suggested by Fleming et al. (2009), it is necessary for the target population to integrate the health messages into routine practice and this can only be done through creation of an enabling environment in which the residents of Luangwa are to make healthier choices for themselves. In as much as this research has few similar results to those of the studies reviewed, it however differs in that it gives a broader review of drivers and barriers to uptake of health messages by the receiving population while suggesting recommendations to improve the implementation process.

The findings of this study revealed various factors which influenced the adoption of health education among the residents of Luangwa district. The simplified messages taught by the health educators made it easy for most people to understand and easily incorporate the preventive measures into routine practice. However, the mode of delivery was not entirely facilitative as some of the recipients would confuse symptoms and modes of transmission of schistosomiasis with those of other diseases such as cholera which were being taught during similar sessions. Secondly, the inclusion of community health workers in delivery of the health messages reduced burden on EHTs who are limited in the district. However, the lack of resources to aid availability of IEC materials faced by health educators proved to take a toll on delivery of the health messages as most suggested it would be more helpful to have visual aids than word of mouth alone. Lastly the various factors associated with facilitating or inhibiting adoption of health education for schistosomiasis were also observed as most barriers were seen to stem from lack of clean water and proper sanitation. Another inhibiting factor was the lack of recognizing schistosomiasis as a problem but as a usual occurrence in the district while most individuals who had been previously exposed to the disease were more prone to adopting the health messages.

It is thus necessary ensure integration of various stakeholders before and during implementation of health education so as ensure its success. This suggests the need to create an enabling environment for the recipients of the health messages from whom behavioral change is expected. An integrated approach for the effective implementation of HE is necessary and can be achieved thorough collaboration with various partners to enable an environment in which people can easily make a decision to adopt the health messages.The availability of clean water and adequate sanitation is particularly necessary to aid uptake of health education and in view of all this, it is necessary for the people of Luangwa to be able to build toilets that cannot easily be washed away or destroyed by the rains and this will restrict urinating and defecating in water sources and the surrounding.

## Recommendations

1. There is need to engage more CHWs as well as monitor & evaluate health education at particular intervals
2. monitor & evaluate the load project of the WASH program to ensure availability of proper toilets so as to provide adequate sanitation for the residents.
3. There is also a need to engage partners who will assist in provision of chlorine in Luangwa and translation of IEC material into local languages. There is need for multi sectorial collaboration between the health & Environmental sector in order to adequately reduce the multiple flooding that occurs in the district as well as with the Agricultural sector to facilitate irrigation systems that will make it more bearable for the residents to participate in farming and thus reduce dependency on fishing.
4. There is a need to have a communication theory generated in future research studies to aid effective implementation of health education and as a result influence it’s uptake.

## Data Availability

All data produced in the present study but not available in the manuscript are available upon request to the authors, however, to the best of our knowledge, all data produced in the present work are contained in the manuscript.

